# Validation and Extension of a Risk Calculator to Predict Mood Recurrence in Young People with Bipolar Disorder

**DOI:** 10.64898/2026.02.20.26346717

**Authors:** Ava Avolio, John Merranko, Mary Kay Gill, Jessica C. Levenson, Tina R. Goldstein, Danella Hafeman, Boris Birmaher

## Abstract

**Objective:** Given the episodic nature of bipolar disorder (BD) and the variability in mood episode recurrence across individuals, accurate recurrence prediction is critical. The original COBY recurrence risk calculator (RC) was developed in a longitudinal youth cohort to estimate threshold recurrence risk. However, its accuracy for predicting subthreshold recurrences had not been fully evaluated. The objective of this study was to extend the previously developed COBY mood recurrence RC to predict both threshold and subthreshold mood recurrences and evaluate its performance in an independent sample.

**Method:** Adolescents and young adults with BD-I/II (N= 51; BD-I: 38, BD-II: 13; 14-24 years old) were interviewed with standard instruments at intake and during the follow-up on average every 6 months for a median of 54 weeks. We assessed the degree to which the COBY RC predicted mood recurrence (threshold or subthreshold) in this independent sample. Discrimination was measured using the area under the receiver operating characteristic curves (AUC); calibration and variable importance were also assessed.

**Results:** The model demonstrated good prediction of any recurrence within the next six months (any threshold recurrence AUC = 0.72, any subthreshold or worse recurrence AUC = 0.77). Calibration analysis demonstrated the model tended to overestimate risk in the external sample, plausibly attributable to differences in recurrence ascertainment strategy (prospective vs retrospective) or the significant difference in prior remission length, a key predictor. Recalibration greatly improved calibration without loss of discrimination.

**Conclusion:** The mood recurrence RC demonstrated good discrimination for both threshold and subthreshold mood recurrences in an independent young adult cohort, consistent with prior youth and adult validations. Validation now spans across developmental stages and different degrees of severity of mood symptoms opening the opportunity for clinical implementation to provide personalized monitoring and early intervention for people with BD.

## Introduction

Bipolar disorder (BD) is a recurrent, episodic illness associated with increased suicidality, substance use, and psychosocial impairment^1–3^. Additionally, mood recurrences contribute to cumulative functional decline, making early intervention imperative^4,5^. Previous study has shown that factors such as early age of onset, symptom severity, number of past recurrences, residual mood symptomatology, and comorbid disorders are associated with elevated risk of mood recurrence^6,7^.

Despite established risk factors and shared diagnostic criteria, the longitudinal course of BD is highly variable between individuals and within clinically defined subtypes (BD-I and BD-II)^8–10^. Prior group-level analyses have shed light regarding different course trajectories and the percentage of follow-up time that people spend in euthymia^11,12^. However, on the individual level, a person may experience one mood episode followed by several years of remission whereas another could experience several full threshold episodes and intermittent residual symptoms each year. This course heterogeneity raises the need for personalized monitoring and treatment because early identification of mood recurrences could enable timely intervention to mitigate cumulative impairment.

To individualize risk assessments, multi-variable models (risk calculators) integrate individual-level data and previously established risk factors to estimate the likelihood of a future event. These tools have been developed and widely use in medicine (e.g., cardiovascular illnesses and cancer)^13–17^. The resulting estimations of risk provide physicians with another reference for decision-making and are not deterministic. Psychiatric risk calculator implementation has been slow compared to other medical disciplines. Of the developed psychiatric risk calculators, focus has mainly targeted psychosis and other high-risk outcomes like suicidality and violence^18–22^. The development of risk calculators (RC) for mood disorders has been sparse, despite their potential value in early identification of mood recurrences in these episodic conditions. Our research group has aimed to address this gap, previously developing three RCs for BD. Within a prediction scope of five years, one RC derived from the Course and Outcome of Bipolar Youth (COBY) study estimates the likelihood of youths previously diagnosed with BP-NOS converting to BD-I or BD-II, while another uses data from the Pittsburgh Bipolar Offspring Study to predict the likelihood children with one or more parents with BD will develop BD themselves^23,24^.

Most recently, the mood recurrence COBY RC was developed to provide individualized threshold mood recurrence predictions for youths, adolescents, and young adults with bipolar spectrum disorders^7^. This model integrated group-level predictors with participants’ longitudinal mood symptom data to produce personalized risk estimates. The RC achieved excellent discrimination with Area Under the ROC Curve (AUC) values ranging from 0.72 to 0.82 for predicting any mood recurrence in a five-year interval, including up to 80% accuracy for depressive episodes and 89% accuracy for hypomanic episodes. Calibration was strong and overall sensitivity and specificity were balanced (0.74), suggesting the model minimized false positives and false negatives. An external validation conducted in an adult cohort (n=258) found the RC predicted recurrence of any threshold episode over a five-year follow up interval with an AUC= 0.77^25^. Together, this model performance, on par with RCs used in other areas of medicine, provides evidence for the model’s generalizability and clinical utility^16,17,21^.

While the original model was an important advance, the RC was trained and tested to only predict full threshold recurrences in those in full remission, constraining its clinical utility. As demonstrated in previous literature, people with BD often experience mood shifts that do not necessarily meet full criteria for a mood episode but represent a meaningful change in subjective experience^26,27^. Despite not reaching the severity of full threshold episodes, these subthreshold symptoms have been associated with functional impairment and increased risk of future threshold mood episodes^28–30^. Training the RC to predict worsening mood offers an opportunity to extend timely care to those in active decline, a group of clear clinical interest. Additionally, the original model evaluated its efficacy on a holdout group from the sample used in construction. The external adult validation provided support for use across the lifespan; however, additional validation in an independently recruited youth and adolescent sample would further establish its robustness for contemporary clinical use.

The current study sought to validate for a third time the previously published COBY threshold mood recurrence RC among youth and young adults with BD. Additionally, utilizing the threshold COBY model as a foundation, we evaluated whether this model could be adapted to predict subthreshold or worse mood recurrences, allowing clinicians to monitor possible mood worsening. External validation and extension of the COBY RC model was conducted using the Predicting Recurrence of Mood in Patients with Bipolar Disorder (PROMPT-BD) cohort, an independent, longitudinally followed sample of youths and young adults diagnosed with BD-I or BD-II. By further externally validating and extending model performance, this study aims to establish the clinical usefulness of the mood recurrence RC for early identification of mood episode recurrences, thus enabling more individualized monitoring and intervention strategies for BD.

## Methods

### Samples

The description of the COBY sample has been previously described in detail^6,30,31^. Briefly, the initial COBY study enrolled 413 youths who met criteria for BD-I, BD-II or Bipolar Disorder Not Otherwise Specified (BD-NOS) as defined by Birmaher et al., 2006. All participants were assessed with the Schedule for Affective Disorders and Schizophrenia for School Age Children-Present and Lifetime Versions (K-SADS-PL) to confirm diagnoses at intake^32^. They were then interviewed at a median of every 7.4 months for a median duration of 12.5 years. At each follow-up, weekly longitudinal psychiatric symptom data were assessed using the Adolescent Longitudinal Interval Follow-up Evaluation (ALIFE)^33^. The ALIFE assigns each week a Psychiatric Status Rating (PSR) score which is a numeric value 1-6 that reflects symptom severity in alignment with DSM-IV criteria^34^. For affective disorders, PSR = 1-2 communicates no or minimal symptoms, 3-4 subthreshold mood symptoms, and 5-6 full threshold moods symptoms.

For the third external validation of the original threshold COBY RC in independent youth and young adult samples, the current study utilized the PROMPT-BD dataset. This prospective cohort study enrolled 51 participants ages 14-25 in outpatient settings from across the United States. At intake, subjects were assessed with either the K-SADS-PL for youths and the Structured Clinical Interview for DSM-5 (SCID-5) for young adults and the ALIFE to evaluate mood fluctuations over the previous three months^35^. For inclusion, participants must have met DSM-5 criteria for BD-I or BD-II (BD-I = 38, BD-II = 13) and had not experienced a full threshold mood episode (PSR ≥ 5) in the previous two months^9,36^. Subjects with current or lifetime diagnoses of psychotic disorders, substance-induced mood disorders, autism, or intellectual disabilities were excluded. PROMPT-BD participants were interviewed using the ALIFE every 6 months and received weekly PSR scores for each of their diagnoses. The PROMPT-BD cohort was recruited after the conclusion of COBY and did not inform the original RC development, allowing for unbiased external validation.

Both the COBY and PROMPT-BD studies were approved by university Institutional Review Boards before enrollment, and informed consent/ assent were obtained from participants and their parents at intake. Likewise, in both studies trained research staff administered semi-structured interview assessments with a ≥ 0.7 interrater reliability and subsequently reviewed cases with a study investigator who assumed final responsibility for all clinical ratings.

### Risk Calculator Development

For inclusion in the original RC development, COBY participants must have achieved full remission (at least two months with minimal to no mood symptoms) and have a history of at least one full-threshold mood episode^7^. The resulting 363 participants (mean age at intake 12.6 ± 3.2 years) were then divided into a training sub-sample (n=182) to develop the RC and a holdout sub-sample (n=181) to assess RC performance. The resulting RC utilized individual ALIFE data and predictor variables previously defined in the literature to estimate risk of full threshold episode recurrences The original COBY model was trained to estimate risk of a threshold mood episode recurrence following remission by using risk associations derived from the construction sample’s ALIFE longitudinal symptom data and a set of predetermined predictor variables. Prediction accuracy in the PROMPT-BD cohort was evaluated by using the ALIFE standardized symptom ratings collected at each PROMPT-BD follow up assessment. A full threshold recurrence was defined as at least one week of a ≥ 5 PSR score for depression, mania, or hypomania during the observation interval.

Using the same calculations as the original model, we extended the RC’s capability by including subthreshold mood recurrences as a secondary outcome. Clinically, a subthreshold mood recurrence was defined as a sustained, meaningful increase in depressive or manic/hypomanic symptoms that did not meet full DSM-V criteria for a mood episode for reasons such as a lack of symptomatology or impairment. A subthreshold or worse recurrence was operationalized as a ≥ 3 PSR score for depression, mania, or hypomania. Subthreshold identification extended the clinical applicability of the original RC by identifying early symptom recurrence that may proceed a full syndromal relapse or functional impairment.

### Statistical Analyses

We trained the RC on the COBY dataset with predictors identified in the existing literature (Birmaher et al., 2020, which included general variables (age, age of mood disorder onset, family history of mania), variables describing the previous mood episode, remission variables, and past episode history variables (See Table 2) ^1,7,11,30,31,37–46^. We trained the model using boosted multinomial classification trees, a useful method for this task because it implicitly fits interactions between predictors. Predictions were then calibrated via Platt scaling^47^. The model was validated on the PROMPT-BD sample and evaluated by estimating area under the receiver operating characteristic curve (AUC) and assessing calibration by plotting observed vs. predicted recurrence risk. Given that the median time until recurrence in PROMPT-BD was 25 weeks, we estimated the six-month risk of recurrence as well as the risk of any recurrence during all available follow-ups. After observing deficiencies in the calibration of model predictions, we compared predictor distributions across the COBY training sample and PROMPT-BD and performed a sensitivity analysis iteratively assigning each individual predictor the value of the mean observed in the COBY training sample and evaluating the impact on model predictions. We further evaluated variable importance in prediction of recurrence risk by estimating the decrement to AUC after sequentially removing each predictor from the model as well as by estimating the mean AUC decrement after permuting predictor values in each test dataset (1000 iterations each). We performed basic sample comparisons using t-tests, chi-squared tests, and Fisher’s exact tests as applicable, and we compared remission durations using scaled Poisson regression and recurrence hazard using Cox proportional hazards regression.

## Results

### PROMPT-BD external validation sample characteristics

The PROMPT-BD external validation sample included 51 participants with a BD-I or BD-II diagnosis who were recruited in remission (mean age = 22.0 ± 2.3, 88% female, 82% white, 75% BD-I). Participants were followed for a median of 54 weeks (ranging 24-106 weeks) with a retention rate of 86%, during which time 45% had a major depressive episode and 33% had a hypo/manic episode (61% had a recurrence of any polarity). Relevant sociodemographic and clinical characteristics for this sample as well as the COBY sample are shown in Table 1.

**Table 1.** Demographic and Clinical Sample Comparisons.

| Variable | COBY (N=363) | PROMPT-BD (N=51) | Test Stat | p-value |
| --- | --- | --- | --- | --- |
| Intake Age | 12.6 (3.2) | 22.0 (2.3) | t=25.69 | **<0.0001** |
| Age at Last Assessment | 25.2 (4.9) | 23.7 (2.1) | t=3.82 | **0.0002** |
| Intake Diagnosis |  |  |  |  |
| Bipolar Disorder-I | 60.1% | 74.5% | $\chi^2=35.27$ | **<0.0001** |
| Bipolar Disorder-II | 6.9% | 25.5% |  |  |
| Bipolar Disorder Not-Otherwise-Specified | 33.1% | 0.0% |  |  |
| Age of Mood Onset | 9.4 (3.9) | 14.6 (3.0) | t=11.25 | **<0.0001** |
| % Female | 47.1% | 88.2% | $\chi^2=30.31$ | **<0.0001** |
| Socioeconomic Status | 4.1 (1.2) | 3.8 (1.0) | t=1.35 | 0.18 |
| Global Assessment of Functioning | 63.6 (14.6) | 74.5 (8.3) | t=7.69 | **<0.0001** |
| Generalized Anxiety Disorder | 39.7% | 62.8% | $\chi^2=9.66$ | **0.002** |
| Attention Deficit Hyperactivity Disorder | 67.3% | 31.4% | 24.78 | **<0.0001** |
| Psychosis | 37.5% | 2.0% | FET | **<0.0001** |
| Substance Use Disorder | 42.2% | 33.3% | $\chi^2=1.55$ | 0.21 |
| Percent who Recurred | 81.0% | 60.8% | $\chi^2=10.38$ | **0.001** |
| Median Remission Length (years) | 2.1 | 1.3 | $\chi^2=24.93$ | **<0.0001** |
| Family History of Bipolar Disorder | 58.1% | 25.5% | $\chi^2=19.44$ | **<0.0001** |
| Family History of Depression | 88.7% | 43.1% | $\chi^2=65.51$ | **<0.0001** |
| Family History of Anxiety | 74.1% | 39.2% | $\chi^2=25.90$ | **<0.0001** |
| Family History of Attention Deficit Hyperactivity Disorder | 46.3% | 21.6% | $\chi^2=11.17$ | **0.0008** |
| Family History of Schizophrenia | 6.9% | 5.9% | FET | ~1 |
| Family History of Substance Use Disorder | 70.0% | 31.4% | $\chi^2=30.05$ | **<0.0001** |

FET = Fisher's Exact Test

### Comparison between PROMPT-BD and COBY sample characteristics

Although PROMPT-BD participants were older on average at recruitment than COBY participants, COBY participants were older at their final assessments used to train the RC than PROMPT-BD participants at their most recent assessments (Table 1). PROMPT-BD participants were also more likely to be female, had higher intake global functioning, were older at mood onset, more likely to have BD-I and generalized anxiety disorder, and less likely to have attention deficit hyperactivity disorder, psychosis, substance use disorder, and family history of all disorders considered except schizophrenia. While a higher percentage of COBY participants had recurrences than PROMPT-BD participants (81% vs. 61%), after accounting for duration of follow-up, PROMPT-BD had a much larger recurrence rate (hazard ratio = 6.04, 95% CI: 3.97-9.17, p<0.0001) and had shorter remission periods (both past and current) on average (incident rate ratio = 0.36, 95% CI: 0.24-0.54, p<0.0001).

### External validation of RC on PROMPT-BD

The threshold RC showed good prediction of any threshold recurrence within the next six months (AUC=0.72) or at any point during the median of 54 weeks of follow-up (time independent AUC=0.74) and performed slightly better when predicting hypo/manic episodes (AUC=0.76) as compared to major depressive episodes (AUC=0.71). As shown in Figure 1, the threshold RC externally validated in PROMPT-BD very comparably to how it internally validated on the COBY holdout sample, with identical AUCs (AUC for both samples=0.72).

**Figure 1.**
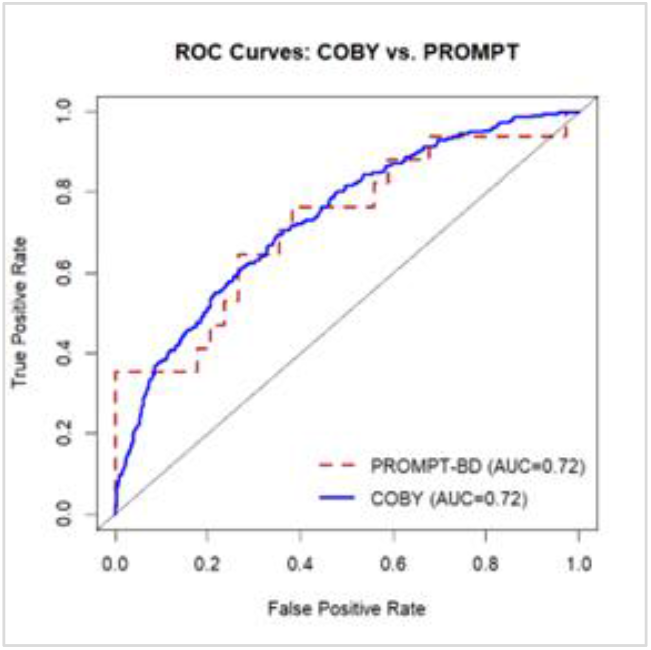

### Calibration of RC predicted risk

The calibration plot shown in Figure 2 indicates that predicted and observed recurrence probabilities were consistent through the range of scores; however, the RC tended to overestimate risk in the PROMPT-BD sample. As previously noted, PROMPT-BD participants’ average durations in remission were significantly shorter than those observed in COBY, and as shown in Table 2, prior remission length was the most influential predictor when training the original RC in COBY. We thus performed a sensitivity analysis iteratively assigning each individual predictor the value of the mean observed in the COBY training sample and evaluating the impact on model predictions. Indeed, results indicated that adjusting prior remission length to take the mean value observed in the COBY training sample had the largest impact on model predictions, reducing the odds of predicting a recurrence by 9%, as compared to -2% to 2% for the other predictor variables. Further, if prior remission length in PROMPT-BD is rescaled to match the mean and variance observed in COBY, calibration improves dramatically (*eFigure 1*).

**Figure 2.**
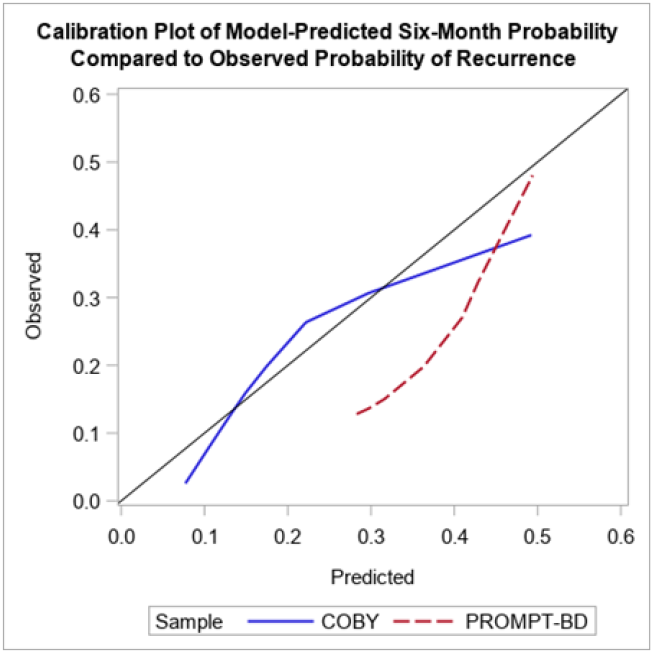

**Table 2.** Variable Importance Statistics.

| Predictor Domain | Predictor Variable | Across-Fold<br>Average<br>Relative<br>Influence | Test AUC Decrement if<br>Removed from Model |  | Test AUC Decrement if<br>Predictor is Permuted |  |
| --- | --- | --- | --- | --- | --- | --- |
|  |  |  | COBY | PROMPT-BD | COBY | PROMPT-BD |
| Participant-Specific<br>Variables | Age (at current period in remission) | 16.23 | 0.025 | 0.008 | 0.026 | 0.005 |
|  | Age of mood disorder onset | 7.31 | 0.016 | 0.060 | -0.012 | 0.045 |
|  | Family History of Mania (1st vs. 2nd vs. Both) | 2.09 | 0.003 | -0.013 | -0.001 | -0.009 |
| Variables From the<br>Previous Episode | Max major depression score | 10.36 | 0.007 | -0.010 | 0.006 | -0.002 |
|  | Number of weeks with threshold major<br>depression | 5.59 | 0.005 | -0.016 | 0.000 | -0.010 |
|  | Max hypo/mania score | 3.63 | 0.000 | -0.005 | 0.002 | -0.011 |
|  | Number of weeks with threshold hypo/mania | 1.56 | 0.000 | 0.010 | -0.002 | 0.002 |
| Remission<br>Variables | Current remission length | 15.84 | 0.009 | 0.037 | 0.040 | 0.046 |
|  | Prior remission length | 31.19 | 0.016 | -0.094 | 0.069 | -0.002 |
| Past Episode<br>History Variables | Number of Recurrences (None vs. 1 vs. 2+) | 0.33 | 0.000 | -0.002 | 0.002 | 0.000 |
|  | Episodes mostly include hypo/manic symptoms? | 5.88 | 0.000 | 0.006 | 0.002 | 0.002 |

### Predictor variable importance

The two most important predictors when predicting risk in PROMPT-BD were age of mood disorder onset and current remission length as evidenced by the largest decrements to AUC being observed after removing those variables from the model or permuting their values (Table 2). The largest disparity in variable importance as estimated on the COBY holdout sample vs. PROMPT-BD was seen in the prior remission length variable, which was the most influential predictor variable when training the RC in COBY and caused the largest AUC decrement if removed or permuted. Conversely, removing or permuting this predictor variable when validating the model in PROMPT-BD improved AUCs, likely due to the large imbalance in the scale of this predictor as previously mentioned.

### Extension of RC to predict subthreshold or worse episodes

When reevaluating the risk calculator to assess its ability to predict six-month risk of subthreshold or worse mood recurrences in PROMPT-BD, the model performed slightly better, with AUCs of 0.77, 0.75, and 0.78 for prediction of any subthreshold or worse mood recurrence, subthreshold or worse major depression, and subthreshold or worse hypo/mania, respectively.

## Discussion

Accurately identifying patients at increased risk for mood recurrences remains a central challenge for the clinical management of BD. This study sought to externally validate the previously developed COBY mood recurrence RC and extend it to predict both threshold and subthreshold mood recurrences. Overall, the COBY RC model maintained good discrimination regardless of outcome (any threshold mood recurrence AUC = 0.72, any subthreshold or worse mood recurrence AUC = 0.77), predicting the next six months in an independent, adolescent and young adult cohort. These performances align with RCs currently utilized in clinical settings^16,17,21^. Several of the most influential predictors in the COBY cohort retained their heightened importance in the PROMPT-BD application, highlighting their continued clinical relevance across developmental stages. The continued strong discrimination between cohorts despite their significant differences in age of mood disorder onset and frequency of mood recurrence demonstrates the model’s ability to be applied to varying populations. Given the unpredictable, episodic nature of BD, a prediction tool that identifies symptom worsening across multiple developmental stages provides a promising resource for clinical responsiveness.

External validations are critical tests of model generalizability communicating its potential to effectively be applied across distinct populations. As compared to COBY, the PROMPT-BD sample tended to develop bipolar disorder later in life and have more frequent mood recurrences (median remission of 25 weeks vs 109 weeks). Despite differences in age of onset and mood recurrence frequency between samples, the RC demonstrated continued discrimination with a moderate calibration shift attributed to these systematic population differences. Notably, this is the third time that the threshold model has demonstrated strong discrimination and the second successful external validation. Another external validation was performed on adults with bipolar in the National Institute of Mental Health Collaborative Depression Study (CDS)^25^. As is the case in the current validation, the CDS validation demonstrated strong discrimination (1-year AUC= 0.77) and a moderate calibration shift. The threshold model tended to underestimate risk in the adult cohort while it tended to overestimate risk in the PROMPT-BD young adult cohort. Given that discrimination remains strong, it is likely that the calibration shift reflects meaningful shifts in sample distribution of important predictor factors. As such, good model performance, despite significant sample variations, supports its transportability with recalibration rather than redevelopment.

Examination of the model’s contributing predictors highlights where it aligns with and where it diverges from previous literature covering recurrence risk. For COBY, the most influential predictors of recurrence were shorter current and previous remission lengths, younger age at time of assessment, younger age of mood disorder onset, and more severe depression symptoms during the previous episode^7^. For the current iteration, permutation analysis shows that age of mood disorder onset and current remission length were most influential. This largely aligns with established literature. Early age of mood onset has been repeatedly identified as a risk factor for more frequent episode recurrence and a higher severity illness course^48,49^. Moreover, current length of remission was one of the most influential factors in both cohorts and has been previously identified as a major risk factor for recurrence in youths and adults^41,44,50^.

Of note, prior remission length was critically important for accurately predicting mood recurrence risk in the original COBY cohort whereas in the PROMPT-BD cohort removing it modestly improved RC performance. A single predictor greatly influencing model performance on both samples but in opposing directions seems contradictory yet reveals a deeper conceptual consistency. Across both cohorts, shortened time since the last episode is a strong indicator of recurrence risk; however, its utility for helping the model distinguish who will relapse first depends on how much participants differ in their remission durations. The COBY sample had less frequent recurrences meaning that the model originally learned to judge time-based risk on longer periods of remission. The PROMPT-BD cohort only recruited individuals with BD-I or BD-II and resulted in a more frequent average recurrence rate, suggesting a higher acuity cohort. Because the PROMPT-BD cohort had recurrence more frequently as a whole, the timeline of recurrence was condensed, giving the entire group a consistently elevated estimated risk. Due to this cohort-wide inflation, current remission length was less helpful in distinguishing who would relapse first. As a result, removing this variable most likely reduced redundancy and allowed for slightly improved discrimination because the model could distinguish between individuals better. Additionally, differences in the observed recurrence rates may reflect ascertainment strategy as mood recurrences were assessed retrospectively in PROMPT-BD but prospectively in COBY. Thus, recall bias may have influenced the observed recurrences rather than true differences in risk.

The model largely aligns with previous mood recurrence findings, but there are also influential risk factors not utilized in the current model. For example, trauma exposure and prior suicide attempts have been associated with more recurrent episodes and ill course^51^. The exclusion of predictive factors in this model is not intended to communicate their insignificance as these factors of suicidal behaviors, non-suicidal self-injury, and psychosis are clinically meaningful and necessary to be addressed by clinicians. Their absence in the model reflects a lack of contribution in predictive power above and beyond the variables already included in the model which may be due to their tendency to highly correlate with the utilized risk predictors.

From a clinical perspective, these results reinforce the need for continuous monitoring and consideration of recent illness course when assessing patient risk. Time since last mood recurrence remains a key signifier of risk especially in settings where remission durations can vary drastically between patients. However, in populations where mood recurrences are more frequent, such as higher acuity or specialized clinics, risk evaluation may rely on behavioral indicators of health rather than time estimations alone. In fact, tools like the current RC should be utilized to inform monitoring intensity, follow-up intervals, and provide anticipatory guidance for patients rather than an attempt to estimate a specific point of recurrence. This focus on risk monitoring provides an exciting opportunity to give patients more responsive care as shortened time to intervention has been shown to be a major contributor to mitigating compounding episode consequences^52,53^. RCs such as the current model would complement stepped-care approaches and alleviate clinician burden as it could provide clinicians guidance to focus pointed monitoring on high-risk patients^54,55^. An important consideration for clinical implementation is the collection of necessary data for model use. The RC requires past episode, demographic, and weekly symptom data. Fortunately, most of these demographic and past episode data are collected during typical clinical intakes. An area of adjustment would be patient symptom monitoring, for which prior work has demonstrated that mood tracking protocols are feasible and clinically informative^56,57^. With three external validations across ages the RC is prepared for clinical implementation. The ALIFE scale has been modified for clinical use and is simple for a trained clinician to administer. The symptom scale and RC are available at https://www.pediatricbipolar.pitt.edu/resources/research-tools.

As with all group-derived prediction models, applying the RC to populations that significantly differ introduces the potential for calibration shift. These models assign individuals risk predictions based on predictor-outcome associations they learn from the construction population. Prior work in multiple medical disciplines has demonstrated that when such models are applied to test populations that meaningfully differ from the construction population, a calibration shift may occur^58,59^. In these shifts, the model can accurately predict which patients have increased risk because the risk associations remain conceptually consistent; however, the degree of risk may not align with the observed outcomes. These shifts do not indicate model failure but instead reflect that the test population has a meaningful difference from the construction population in a key predictor. Site-specific recalibration has been shown to maintain RC clinical effectiveness by allowing it to respond to distinct clinical populations^60,61^. As in the case with the current model, when the distribution of prior remission length in the PROMPT-BD sample was shifted to reflect the distribution in the COBY sample, the calibration significantly improved (*see eFigure 1*). From this perspective, recalibration can be considered a feature because it allows the same tool to be effectively applied across varying clinical settings such as acuity level or age of patient population served. That said, recalibration may not be universally required. Emerging strategies propose comparing the alignment of historical trends in the target population and original construction cohort or observing performance during a base model trial before determining if recalibration is necessary^21,59,60^.

A future opportunity to build the robustness of the current mood recurrence RC would be to pair this group-level RC with a more individualized, dynamic one. In this approach, the current RC would serve as a foundation for individuals’ personal data to inform. Incorporating on-going longitudinal data would allow the RC to update its risk estimates as new individual mood trends emerge. Dynamic updating alleviates the reliance on fixed baseline predictors, thereby reducing the need for site specific recalibration. Emerging passive sensing data collection tools, such as sleep-monitoring smart watches and smartphone applications, may offer the opportunity to personalize prediction models that evolve with the illness course, treatment adjustments, and environmental exposures of each patient^62–64^. Although personalized dynamic models represent an exciting future direction, their success is dependent upon the validation and generalizability of established RCs such as the present model.

These results and their application to clinical settings should be viewed in context of the study’s limitations. First the RC was derived from and externally validated on data from participants who were majority white, female, and recruited from out-patient settings which may limit generalizability to populations with different demographics or clinical profiles without adjustment. Second, COBY and PROMPT-BD are both longitudinally followed cohorts, but symptomatology was collected retrospectively using structured research instruments. These assessments may be vulnerable to recall bias and may differ from the symptom collection procedures in routine clinical practice. Additionally, reliance on research assessments may not fully capture the factors influencing recurrence risk in real world settings such as treatment adherence and environmental stressors.

In conclusion, the present study successfully extended and validated a mood recurrence RC to predict threshold and subthreshold or worse bipolar mood episodes. The predictive accuracies for six-month predictions are within accepted limits and consistent with tools previously adopted by other medical disciplines. This study represents a third validation of the threshold recurrence RC and a validation of a subthreshold or worse prediction extension, further affirming the tool’s ability to be clinically utilized. Together, these findings support the growing role of RCs in advancing precision psychiatry and data-informed care. Future work should focus on integration of longitudinal passive sensing data into dynamic models and prospective implementation studies to evaluate clinical impact. The RC and symptoms scales are free for clinical use and can be found at https://www.pediatricbipolar.pitt.edu/resources/research-tools.

## Data Availability

All data produced in the present study are available upon reasonable request to the authors.

---

**Figure e1.**
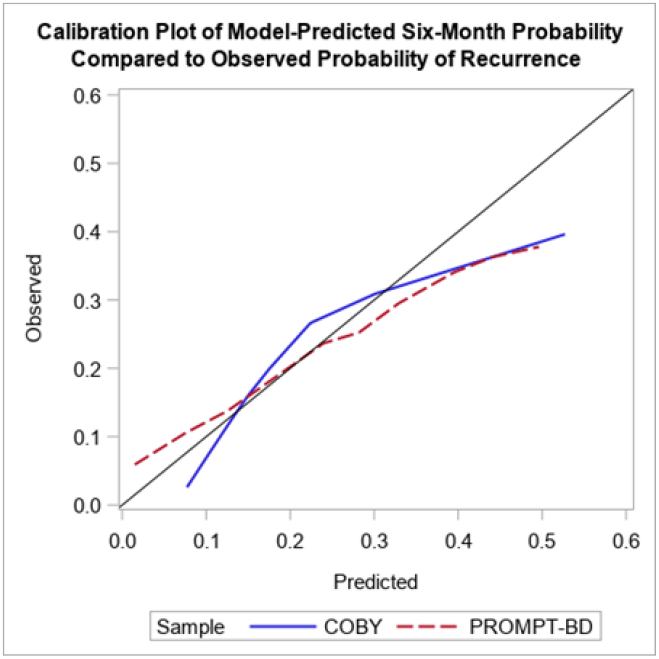

